# Who defines the “personal utility” of genetic and genomic testing? A systematic review

**DOI:** 10.1101/2022.12.04.22283084

**Authors:** Emily G. Miller, Jennifer L. Young, Anoushka Rao, Eliana Ward-Lev, Meghan C. Halley

## Abstract

**Importance:** Expansion in the clinical use of genetic and genomic testing has led to a recognition that these tests provide personal as well as clinical utility to patients and families. It is essential to ensure that members of diverse sociodemographic backgrounds are included in defining and measuring personal utility.

**Objective:** To determine the demographics of participants contributing to the development of a definition of personal utility for genetic and genomic testing.

**Evidence Review:** We searched PubMed, Scopus, Web of Science, and Embase for peer-reviewed literature published between 2003 and January 2022 on the personal utility of genetic or genomic sequencing. Our review included both qualitative and quantitative studies with samples that included patients, their family members, or the general public. Eligible studies could examine any clinical genetic or genomic test and required the use of the term “utility.” Authors extracted and reviewed study and participant characteristics including number of participants, study location (U.S. or international), primary methodology (qualitative or quantitative), race and ethnicity, gender, income and education data.

**Findings:** Our final review included 53 studies and 13,315 total participants. Gender was provided for 95.6% of participants (n=12,724), of whom 61.5% were female (n=7,823). Race and/or ethnicity was provided for 83.0% of participants (n=11,048), of whom 82.2% (n=9,083) were White. The remaining participants were identified as Hispanic/Latinx (5.5%, n=607), Asian American and Pacific Islander (3.8%, n=421), Black (3.5%, n=387), multiracial (0.2%, n=27), and various other racial or ethnic categories (3.7%, n=412). Educational attainment was reported for 82.6% of participants. Among these participants, 71.2% (n=7,830) had a bachelor’s degree or higher. Income was reported for 66.5% of participants (n=8,857), and 66% of these participants (n=5,831) reported income above the U.S. median.

**Conclusions and Relevance:** Our results suggest that the concept of personal utility in genetic and genomic testing in the U.S. is disproportionately defined by the perspectives a narrow subset of the population – specifically non-Hispanic White, well-educated women with above-average household incomes. If we are to provide equitable care in the areas of genomics and genetics, we will need to expand research to include more diverse and representative samples.

**Key Points:** *Question:* What are the demographic characteristics of participants contributing to a definition of personal utility in genetic and genomic testing?

*Findings:* This systematic review included 53 studies and13,315 total participants. Demographic characteristics on gender were provided for 95.6% of participants, race and/or ethnicity for 83.0% of participants, educational attainment for 82.6% of participants, and income for 66.5% of participants. Participants were 61.5% female, 82.2% White, 71.2% had a bachelor’s degree or higher, and 66% reported income above the U.S. median.

*Meaning:* Our understanding of the utility of genetics and genomics lacks the perspectives of people with diverse sociodemographic characteristics.

## Introduction

As clinical use of genetic and genomic testing grows, clinicians and researchers are increasingly recognizing that, beyond clinical utility, these tests provide substantial personal utility to patients and families.^1^ In response, researchers have employed diverse methods to define and measure the broad range of subjective outcomes associated with personal utility in this context. Key dimensions of personal utility have captured psychosocial benefits, facilitated social support, improved care coordination, and information for reproductive and other healthcare decision-making.^1,2^

It is essential to ensure that the voices of patients, caregivers from diverse sociodemographic backgrounds are included in defining and measuring personal utility. While the need to diversify genomic datasets has been widely recognized,^3^ the parallel need to include diverse perspectives when defining the value of genomic data for patients and families remains unexplored. Recent systematic reviews on this topic have not reported the sociodemographic makeup of participants included in the reviewed studies.^1,2^ To address this gap, we conducted a systematic review of peer-reviewed research focused on defining and measuring the personal utility of genetic and genomic testing to determine the underlying sociodemographic characteristics of study participants.

## Materials and Methods

Our systematic review utilized and updated a widely-cited 2017 systematic review by Kohler et al on the personal utility of genetic and genomic testing.^1^ We updated Kohler et al’s original search string to account for changing terminology over time (e.g., including “perceived” as well as “personal” utility) and conducted a new search for studies subsequently published in the same databases. Additional methodological information and the full details of our search string are provided in **Supplementary Appendix 1**.

Studies identified were screened for eligibility by two independent reviewers. Eligibility criteria included:

1. Focused on perspectives of patients, their family members, or the general public;
2. Focused on any clinical genetic or genomic test;
3. Provided empirical data on participants’ perspectives of the “utility” of genetic or genomic tests;
4. Published between August 2016 (the date of Kohler et al’s final search) and January 2022;
5. Published in English in a full-text form;
6. Conducted in the United States.

## Data Extraction and Analysis

Three authors extracted and reviewed study and participant characteristics including: number of participants, study location (U.S. or international), primary methodology (qualitative or quantitative), race and ethnicity, gender, income, and education data. Methods for harmonizing sociodemographic categories are provided in **Supplementary Appendix 1**, and the complete data dictionary is provided in **Supplementary Appendix 2**. We consolidated individual-level data across all studies for descriptive analyses and used a Pearson’s chi-squared test to compare demographic characteristics by study methodology, with a threshold of p<0.05 for statistical significance. The complete analytic dataset is provided in **Supplementary Appendix 3**.

## Results

Our final review included 53 eligible studies with 13,315 total participants (**Figure 1**). Of these, 32 studies (60.4%) used primarily qualitative methodology, although these accounted for only 14.1% (n=1,877) of all participants. A descriptive overview is provided in **Figure 2**, and a summary of comparisons in **Table 1**. Additional descriptive details are provided in **Supplementary Appendix 4**.

**Figure 1:**
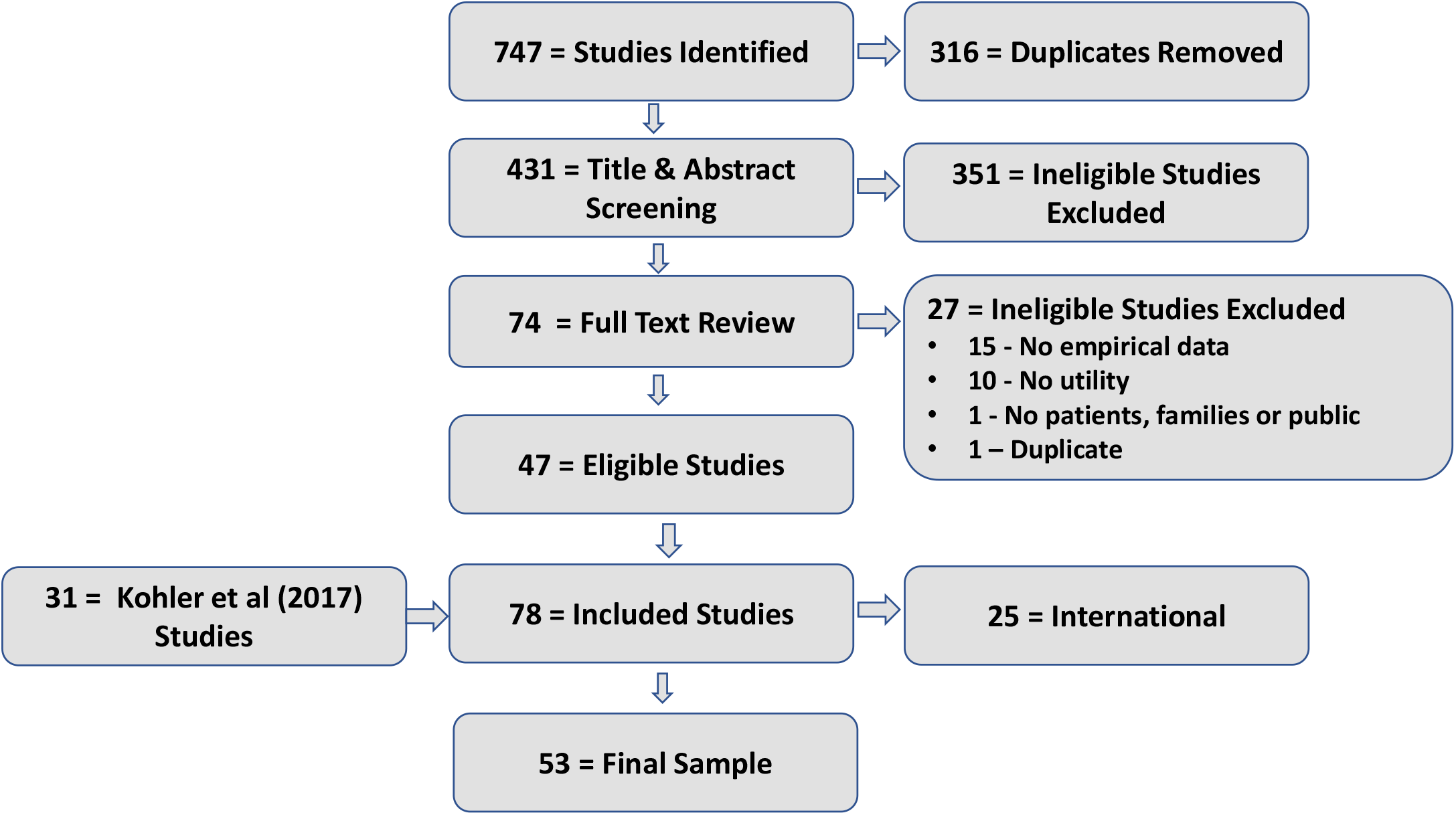
PRISMA Diagram.

**Figure 2:**
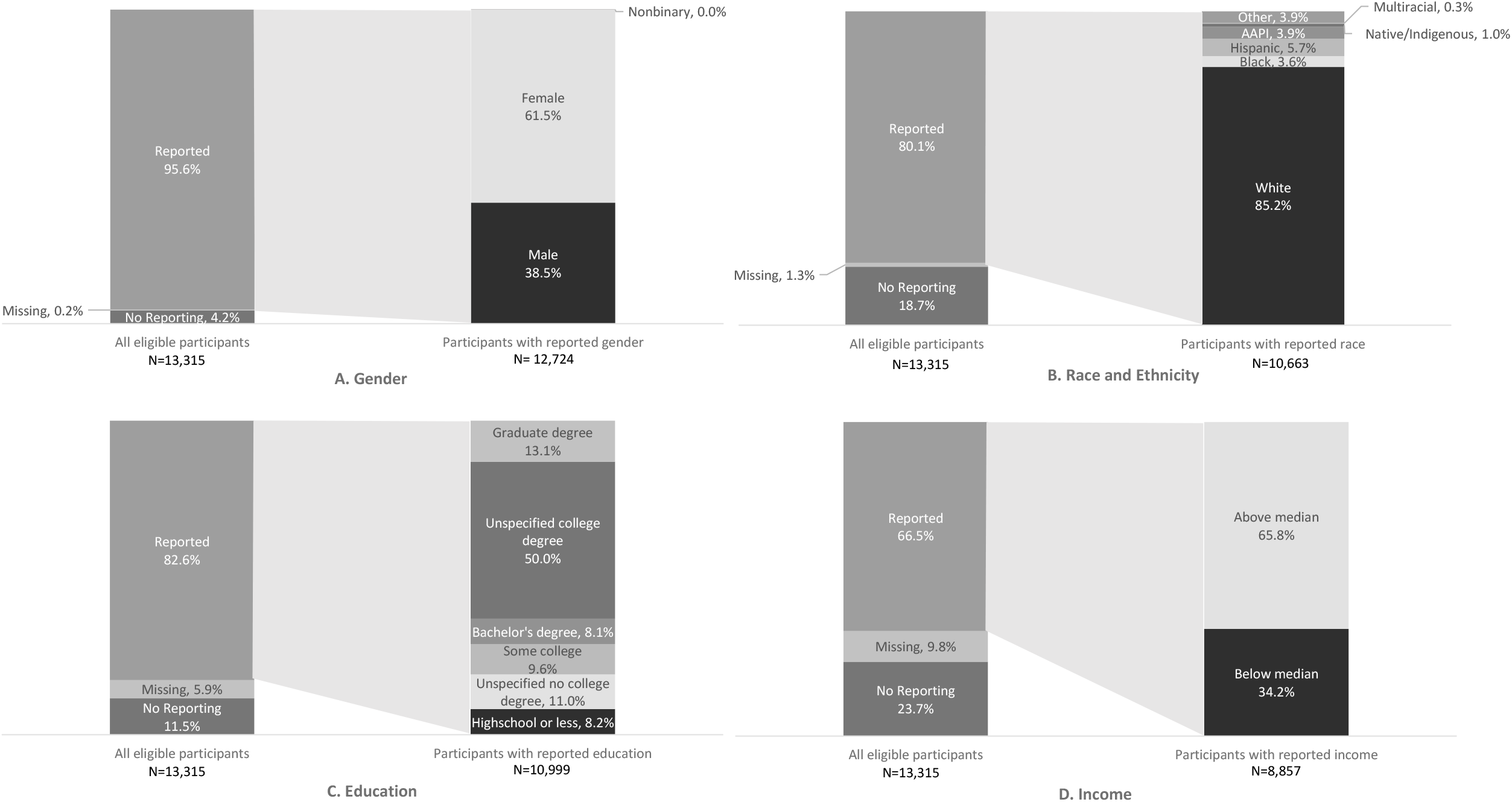
Comparison of demographics and reporting across studies.

**Table 1:** Comparison of demographics in primarily quantitative versus primarily qualitative studies.

| | All eligible<br>n(%) | Quantitative<br>n(%) | Qualitative<br>n(%) | Chi-Squared Test<br>$\chi^2(df)$, p-value |
| --- | --- | --- | --- | --- |
| **Gender** |  |  |  |  |
| Women | 7,823 (61.5%) | 6,573 (60.2%) | 1,229 (69.7%) | $\chi^2(1)=57.2$<br>$p<0.0001^{***}$ |
| Men | 4,898 (38.5%) | 4,341 (39.8%) | 535 (30.3%) |  |
| Total reported* | 12,724 (100%) | 10,917 (100%) | 1,764 (100%) |  |
| **Race & Ethnicity\*\*** |  |  |  |  |
| White | 9,083(85.2%) | 7,950(87.7%) | 1,109(71.0%) | $\chi^2(1)=162.2$<br>$p<0.0001^{***}$ |
| Any Other Race/Ethnicity | 1,965(18.4%) | 1,494(16.5%) | 455(29.1%) |  |
| Total reported | 10,663(100%) | 9,061(100%) | 1,562(100%) |  |
| **Education** |  |  |  |  |
| College degree or more | 7,830 (71.2%) | 6,977 (71.4%) | 826 (69.9%) | $\chi^2(1)=1.0$<br>$p=0.311$ |
| No college degree | 3,169 (28.8%) | 2,801 (28.6%) | 355 (30.1%) |  |
| Total reported | 10,999 (100%) | 9,778 (100%) | 1,181 (100%) |  |
| **Income** |  |  |  |  |
| Below Median | 3,026 (34.2%) | 2,764 (35.0%) | 256 (28.2%) | $\chi^2(1)=16.7$<br>$p<0.0001^{***}$ |
| Above Median | 5,831 (65.8%) | 5,144 (65.0%) | 653 (71.8%) |  |
| Total reported | 8,857 (100%) | 7,908 (100%) | 909 (100%) |  |

\*Also includes 3 nonbinary respondents
\*\*Participants could select more than one category; numbers in each category exceed count for total reported
\*\*\*Significant at $p<0.05$

### Gender

Gender was the most frequently reported demographic variable, provided for 95.6% of participants (n=12,724). Only two studies included participants reporting a gender other than male or female (n=3 participants), and 11 studies reported only one gender, implying binary gender categories. Women made up 61.5% (n=7,823) of all participants, and only slightly less (59.7%) when the seven studies of only women were excluded. A greater proportion of qualitative study participants were women (69.7%, n=1,229) compared to quantitative study participants (60.2%, n=6,573), a difference that was statistically significant (*X*^*2*^ = 57.2, *p* <0.0001).

### Race and Ethnicity

Race and/or ethnicity were provided for 80.1% of participants (n=10,663). Six studies allowed participants to choose more than one category, therefore percentages sum to more than 100%. The vast majority (85.2%, n=9,083) identified as White. The remaining participants included Hispanic/Latinx (5.7%, n=607), Asian American and Pacific Islander (3.9%, n=421), Black (3.6%, n=387), multiracial (0.3%, n=27), and various other racial or ethnic categories (3.9%, n=412). The percentage of participants identifying as White was higher in quantitative studies (84.2%, n=7,950) than in in qualitative studies (70.9%, n=1,109), a difference which was statistically significant (*X*^*2*^ = 162.2, *p* <0.0001).

### Education

Educational attainment was reported for 82.6% of participants (n=10,999). Among these participants, 71.2% (n=7,830) had at least a bachelor’s degree, compared to 28.8% (n=3,169) without. Further, among the 2,333 participants who specified their level of college degree, 61.7% (n=1,440) had a graduate degree (**Supplementary Appendix 4**). The proportion of participants reporting any college degree in qualitative and quantitative studies was not statistically significant.

### Income

Income was reported for 66.5% of participants (n=8,857). Of these, 65.8% (n=5,831) reported income above the U.S. median, while 34.2% (3,026) reported an income below the median. An even greater proportion of qualitative study participants reported income above the median (71.8%, n=653) compared to quantitative study participants (65.0%, n=5,144), a difference that was statistically significant (*X*^*2*^ = 16.7, *p* <0.0001).

## Discussion

Our results suggest that the literature on the personal utility of genetic and genomic testing in the United States has disproportionately captured the perspectives of a narrow subset of the population – specifically White, well-educated women with above-average household incomes. These findings are particularly concerning given recent research suggesting important differences in the perceived utility of genetic and genomic tests across participants with diverse backgrounds.^5^ These data also are not representative of the U.S. population. In the U.S., only 37.9% of adults have a Bachelor’s degree, only 61% identify as White, approximately half are women and, of course, 50% are below the median income.

In terms of gender and income, these trends appear even more pronounced in qualitative studies of utility, which is concerning given the foundational role these types of studies play in defining the concept of personal utility. In addition, our review identified only 274 Black participants in any eligible quantitative study of utility, compared to 7,950 non-Hispanic White participants, and nearly half (n=41) of all Black participants in qualitative studies came from a single study specifically focused the perspectives of Black patients.^6^ Interestingly, although many other studies reviewed include all or nearly all White participants, these studies don’t state an explicit focus on understanding the perspectives of White patients. Such patterns only serve to normalize and reinforce narratives prioritizing White perspectives.

Further, information on education and income are either not reported or missing for up to 35% of all study participants. Given the potential for such sociodemographic characteristics to influence the personal utility of genetics and genomics, future studies should ensure robust and systematic collection of participants’ sociodemographic characteristics.

## Conclusion

The results of our systematic review suggest that our understanding of the utility of genetics and genomics is disproportionately based on the perspectives of White women with high income and education. In order to achieve the National Human Genome Research Institute’s goal of “equitable use of genomics in healthcare that avoids exacerbating and strives towards reducing health disparities,”^7^ we need not only to collect diverse genomic data but also to understand perspectives on the value of these data for diverse patients and families.

## Data Availability

All data produced in the present work are contained in the manuscript

## Acknowledgements

The authors would like to thank Charis Tang for her assistance in manuscript preparation.

---

**Supplementary Appendix 3:** Data Dictionary.

| Variable Name | Variable Type | Definition | Codes |
| --- | --- | --- | --- |
| UPI | Text | Assigned unique identifier |  |
| DOI | Text | DOI from PubMed |  |
| Title | Text | Study Title |  |
| Journal | Text | Journal Name or Abbreviation |  |
| Year | Text | Year of Publication |  |
| N_Study | Numeric | Total number of all types of participants in the study. |  |
| N_Patient | Numeric | Total number of individuals identified as patients in the study. |  |
| N_Family | Numeric | Total number of individuals identified as family members of patients or caregivers in the study. Include here "advocates" or other individuals whose opinions are collected as a proxy for patients. Do not include individuals recruited as members of the general public who happen to be parents. |  |
| N_Public | Numeric | Total number of individuals identified as members of the general public. |  |
| N_Ineligible | Numeric | Total number of individuals in the study who are not eligible (e.g., physicians, policymakers) |  |
| N_Eligible | Numeric | Total number of patients, family members, or public from whom data on utility was collected in the study. |  |
| Method_Type | Categorical | Indicates whether the paper primarily reporting on a quantitative or a qualitative method. Qualitative papers tend to be exploratory, small samples and involve interviews, focus groups, or case studies. Quantitative papers tend to use surveys and have larger sample sizes. If both methods were used, primary method determined by consensus. For example, a study that includes a sample where all are interviewed and participants also complete a survey is characterized as qualitative. If all participants complete a survey and only a small subset are interviewed, then it is characterized as a survey. | Qual = Qualitative;<br>Quant = Quantitative |
| US_Int | Categorical | Indicates whether the participants in the study from the United States only or including (or only) international participants. | US Only vs.<br>International |
| Edu_Rep | Binary | Indicates whether the paper presents data on educational attainment for eligible participants. | Yes/No |
| N_HS_OrLess | Numeric | Number of participants categorized as having completed a high school degree or less. See Supplementary Appendix 2 for details |  |
| N_Some_College | Numeric | Number of participants categorized as having completing education beyond highschool but not completed a four-year college degree. See Supplementary Appendix 2 for details |  |
| N_LessThan_CollegeDeg | Numeric | Number of participants categorized as having not completed a four-year college degree. See Supplementary Appendix 2 for details. Only use if data not disaggregated by N_HS_OrLess and N_Some_College. |  |
| N_CollegeDeg_OrMore | Numeric | Number of participants categorized as having completed a four-year college degree or more education. See Supplementary Appendix 2 for details. Only use if data not disaggregated by N_Bachelor_Deg and Graduate_Deg. |  |
| N_Bachelor_Deg | Numeric | Number of participants categorized as having completed a four-year college degree. See Supplementary Appendix 2 for details. |  |
| N_Graduate_Deg | Numeric | Number of participants categorized as having completed a graduate degree. See Supplementary Appendix 2 for details. |  |
| N_Edu_Miss | Numeric | Number of participants for whom education data is missing, even though study does report education in general. Includes missing, not reported, or declined. Also includes undefined "other" category if used. |  |
| N_Edu_Rep_Total | Numeric | Total number of individuals with reported education data (does not include those with missing data). |  |
| N_Inc_Rep | Binary | Indicates whether the paper reports data on income for eligible participants. | Yes/No |
| N_Inc_Miss | Numeric | Number of participants for whom income data is missing, even though study does report income in general. Includes missing, not reported, or declined. |  |
| N_Inc_BelowMedian | Numeric | Number of participants with income below the U.S. median. Calculated using US population income distribution. See Supplementary Appendix 1 for details |  |
| N_Inc_AboveMedian | Numeric | Number of participants with income below the U.S. median. Calculated using US population income distribution. See Supplementary Appendix 1 for details |  |
| N_Inc_Rep | Numeric | Total number of participants for with income data reported (does not include those with missing data). |  |
| Gen_Rep | Binary | Indicates whether the paper reports data on gender for eligible participants. | Yes/No |
| Gen_Women_Only | Binary | Indicates whether the paper focuses on (and only recruit) women. | Yes/No |
| N_Women | Numeric | Number of participants categorized as women. See Supplementary Appendix 2 for details |  |
| N_Men | Numeric | Number of participants categorized as men. See Supplementary Appendix 2 for details |  |
| N_Gen_Other | Numeric | Number of participants self-reporting a gender other than man or woman. See Supplementary Appendix 2 for details |  |
| N_Gen_Miss | Numeric | Number of participants for whom gender data is missing. Includes missing, not reported, or declined. |  |
| N_Gen_Rep_Total | Numeric | Total number of individuals with reported gender data (does not include those with missing data). |  |
| Race_Rep | Binary | Indicates whether the paper reports race and/or ethnicity data. Do not treat country of origin as race/ethnicity if this is the only information provided. If race or ethnicity is only reported for a portion of the sample (e.g., only reports % White ) note in Race_Rep_Notes. | Yes/No |
| Race_Rep_Notes | Text | If race and/or ethnicity are only reported for some only some categories, please explain here. |  |
| Race_Focus | Binary | Indicates whether the study explicitly states a focus (and only recruit) individuals from a particular race or ethnicity. | Yes/No |
| N_NHWhite | Numeric | Number of participants categorized as non-Hispanic White. See Supplementary Appendix 2 for details |  |
| N_Black | Numeric | Number of participants categorized as Black. See Supplementary Appendix 2 for details |  |
| N_HispLatx | Numeric | Number of participants categorized as Hispanic or Latinx. See Supplementary Appendix 2 for details |  |
| N_AAPI | Numeric | Number of participants categorized as Asian-American or Pacific Islander. See Supplementary Appendix 2 for details |  |
| N_Native_Indig | Numeric | Number of participants categorized as Native American or another indigenous group. See Supplementary Appendix 2 for details |  |
| N_Multiracial | Numeric | Number of participants categorized as multiracial. See Supplementary Appendix 2 for details |  |
| N_Race_Other | Numeric | Number of participants categorized as another race or ethnicity. See Supplementary Appendix 2 for details |  |
| N_Race_Miss | Numeric | Number of participants for whom race and ethnicity data is reported as missing, even though study does report this information in general. Includes missing, not reported, or declined. |  |
| N_Race_Rep_Total | Numeric | Total number of individuals with reported race and/or ethnicity data (does not include those with missing data). |  |
| Race_Discrete | Binary | Indicates a study in which individuals were allowed to select multiple options for race and/or ethnicity. | Yes/No |

