## Supplementary Appendix 1 for "Who defines the “personal utility” of genetic and genomic testing? A systematic review"

### Supplementary Appendix 1: Additional Methodological Detail

#### Part A: Search Strategy

Following Kohler et al's original methodology, we utilized two complementary search strings to query each of the following databases: PubMed, Scopus, Web of Science, and Embase. For each database, the first search string was designed to search both titles and abstracts for terms related to genetic testing combined with terms related to personal utility. The second string used a broader search for personal utility. Instead of phrase searches, it constructed the search into personal/patient/individual terms combined with utility/value/meaning terms (for example, it searched (personal AND utility) rather than “personal utility”) These searches were only for keywords in the title field. This strategy was designed to identify relevant articles that did not use the exact phrases for personal utility without adding excessive false positives to the search results.

The search strings used were as follows:

***PubMed***

("genetic testing"[majr] OR "genetic testing"[tiab] OR "genetics testing"[tiab] OR "genome testing"[tiab] OR "genomic testing"[tiab] OR "genomics testing"[tiab] OR "genetic sequencing"[tiab] OR "genetics sequencing"[tiab] OR "genome sequencing"[tiab] OR "genomic sequencing"[tiab] OR "genomics sequencing"[tiab] OR "exome sequencing"[tiab] OR "exomic sequencing"[tiab] OR "sequence analysis, dna"[majr] OR "genetic profiling"[tiab] OR "genome profiling"[tiab] OR "genomic profiling"[tiab] OR "genomics profiling"[tiab] OR "direct to consumer testing"[tiab] OR "direct to consumer genetic"[tiab] OR "direct to consumer genetics"[tiab] OR "direct to consumer genome"[tiab] OR "direct to consumer genomic"[tiab] OR "direct to consumer genomics"[tiab] OR "personal genetic"[tiab] OR "personal genetics"[tiab] OR "personal genome"[tiab] OR "personal genomic"[tiab] OR "personal genomics"[tiab] OR "individual genetic"[tiab] OR "individual genetics"[tiab] OR "individual genome"[tiab] OR "individual genomic"[tiab] OR "individual genomics"[tiab] OR "genetic susceptibility testing"[tiab] OR "genomic risk profiling"[tiab] OR "genome based testing"[tiab]) AND ("personal utility"[tiab] OR "patient utility"[tiab] OR "individual utility"[tiab] OR "participant utility"[tiab] OR "consumer utility"[tiab] OR "public utility"[tiab] OR "health utility"[tiab] OR "health-related utility"[tiab] OR “perceived utility”[tiab] OR “patient-oriented utility”[tiab] OR "personal meaning"[tiab] OR "patient meaning"[tiab] OR "individual meaning"[tiab] OR "participant meaning"[tiab] OR "consumer meaning"[tiab] OR "public meaning"[tiab] OR "personal value"[tiab] OR "patient value"[tiab] OR "individual value"[tiab] OR "participant value"[tiab] OR "consumer value"[tiab] OR "public value"[tiab] OR "personal benefit"[tiab] OR "personal benefits"[tiab] OR "personal harm"[tiab] OR "personal harms"[tiab] OR "personal outcome"[tiab] OR "personal outcomes"[tiab] OR "non-clinical benefit"[tiab] OR "non-clinical benefits"[tiab] OR "nonclinical benefit"[tiab] OR "nonclinical benefits"[tiab] OR "non-medical benefit"[tiab] OR "non-medical benefits"[tiab] OR "nonmedical benefit"[tiab] OR "nonmedical benefits"[tiab] OR “psychological outcome”[tiab] OR “psychological impact”[tiab] OR “psychological outcomes”[tiab] OR “psychological impacts”[tiab])

("genetic testing"[majr] OR "genetic testing"[tiab] OR "genetics testing"[tiab] OR "genome testing"[tiab] OR "genomic testing"[tiab] OR "genomics testing"[tiab] OR "genetic sequencing"[tiab] OR "genetics sequencing"[tiab] OR "genome sequencing"[tiab] OR "genomic sequencing"[tiab] OR "genomics sequencing"[tiab] OR "exome sequencing"[tiab] OR "exomic sequencing"[tiab] OR "sequence analysis, dna"[majr] OR "genetic profiling"[tiab] OR "genome profiling"[tiab] OR "genomic profiling"[tiab] OR "genomics profiling"[tiab] OR "direct to consumer testing"[tiab] OR "direct to consumer genetic"[tiab] OR "direct to consumer genetics"[tiab] OR "direct to consumer genome"[tiab] OR "direct to consumer genomic"[tiab] OR "direct to consumer genomics"[tiab] OR "personal genetic"[tiab] OR "personal genetics"[tiab] OR "personal genome"[tiab] OR "personal genomic"[tiab] OR "personal genomics"[tiab] OR "individual genetic"[tiab] OR "individual genetics"[tiab] OR "individual genome"[tiab] OR "individual genomic"[tiab] OR "individual genomics"[tiab] OR "genetic susceptibility testing"[tiab] OR "genomic risk profiling"[tiab] OR "genome based testing"[tiab]) AND ((personal[ti] OR patient[ti] OR individual[ti] OR participant[ti] OR consumer[ti]) AND (utility[ti] OR meaning[ti] OR value[ti] OR benefit[ti])))

***Scopus***

TITLE ( "genetic testing" OR "genetics testing" OR "genome testing" OR "genomic testing" OR "genomics testing" OR "genetic sequencing" OR "genetics sequencing" OR "genome sequencing" OR "genomic sequencing" OR "genomics sequencing" OR "exome sequencing" OR "exomic sequencing" OR "sequence analysis" OR "genetic profiling" OR "genome profiling" OR "genomic profiling" OR "genomics profiling" OR "direct to consumer testing" OR "direct to consumer genetic" OR "direct to consumer genetics" OR "direct to consumer genome" OR "direct to consumer genomic" OR "direct to consumer genomics" OR "personal genetic" OR "personal genetics" OR "personal genome" OR "personal genomic" OR "personal genomics" OR "individual genetic" OR "individual genetics" OR "individual genome" OR "individual genomic" OR "individual genomics" OR "genetic susceptibility testing" OR "genomic risk profiling" OR "genome based testing" ) AND TITLE-ABS-KEY ( "personal utility" OR "patient utility" OR "individual utility" OR "participant utility" OR "consumer utility" OR "public utility" OR "health utility" OR "health-related utility" OR “perceived utility” OR “patient-oriented utility” OR "personal meaning" OR "patient meaning" OR "individual meaning" OR "participant meaning" OR "consumer meaning" OR "public meaning" OR "clinical meaning" OR "medical meaning" OR "personal value" OR "patient value" OR "individual value" OR "participant value" OR "consumer value" OR "public value" OR “psychological outcome” OR “psychological impact” OR “psychological outcomes” OR “psychological impacts” OR "personal benefit" OR "personal benefits" OR "personal harm" OR "personal harms" OR "personal outcome" OR "personal outcomes" OR "non-clinical benefit" OR "non-clinical benefits" OR "nonclinical benefit" OR "nonclinical benefits" OR "non-medical benefit" OR "non-medical benefits" OR "nonmedical benefit" OR "nonmedical benefits" ) AND ( LIMIT-TO ( LANGUAGE , "English" ) )

( TITLE ( "genetic testing" OR "genetics testing" OR "genome testing" OR "genomic testing" OR "genomics testing" OR "genetic sequencing" OR "genetics sequencing" OR "genome sequencing" OR "genomic sequencing" OR "genomics sequencing" OR "exome sequencing" OR "exomic sequencing" OR "sequence analysis" OR "genetic profiling" OR "genome profiling" OR "genomic profiling" OR "genomics profiling" OR "direct to consumer testing" OR "direct to consumer genetic" OR "direct to consumer genetics" OR "direct to consumer genome" OR "direct to consumer genomic" OR "direct to consumer genomics" OR "personal genetic" OR "personal genetics" OR "personal genome" OR "personal genomic" OR "personal genomics" OR "individual genetic" OR "individual genetics" OR "individual genome" OR "individual genomic" OR "individual genomics" OR "genetic susceptibility testing" OR "genomic risk profiling" OR "genome based testing" ) ) AND ( TITLE ( personal OR patient OR individual OR participant OR c onsumer ) AND TITLE ( utility OR meaning OR value OR benefit ) )

***Web of Science***

TITLE: ("genetic testing" OR "genetics testing" OR "genome testing" OR "genomic testing" OR "genomics testing" OR "genetic sequencing" OR "genetics sequencing" OR "genome sequencing" OR "genomic sequencing" OR "genomics sequencing" OR "exome sequencing" OR "exomic sequencing" OR "sequence analysis" OR "genetic profiling" OR "genome profiling" OR "genomic profiling" OR "genomics profiling" OR "direct to consumer testing" OR "direct to consumer genetic" OR "direct to consumer genetics" OR "direct to consumer genome" OR "direct to consumer genomic" OR "direct to consumer genomics" OR "personal genetic" OR "personal genetics" OR "personal genome" OR "personal genomic" OR "personal genomics" OR "individual genetic" OR "individual genetics" OR "individual genome" OR "individual genomic" OR "individual genomics" OR "genetic susceptibility testing" OR "genomic risk profiling" OR "genome based testing") *AND* TOPIC: ("personal utility" OR "patient utility" OR "individual utility" OR "participant utility" OR "consumer utility" OR "public utility" OR "health utility" OR "health-related utility" OR “perceived utility” OR “patient-oriented utility” OR "personal meaning" OR "patient meaning" OR "individual meaning" OR "participant meaning" OR "consumer meaning" OR "public meaning" OR “psychological outcome” OR “psychological impact” OR “psychological outcomes” OR “psychological impacts” OR "personal value" OR "patient value" OR "individual value" OR "participant value" OR "consumer value" OR "public value" OR "clinical value" OR "medical value" OR "personal benefit" OR "personal benefits" OR "personal harm" OR "personal harms" OR "personal outcome" OR "personal outcomes" OR "non-clinical benefit" OR "non-clinical benefits" OR "nonclinical benefit" OR "nonclinical benefits" OR "non-medical benefit" OR "non-medical benefits" OR "nonmedical benefit" OR "nonmedical benefits")
Refined by:LANGUAGES: ( ENGLISH )

TITLE: ("genetic testing" OR "genetics testing" OR "genome testing" OR "genomic

testing" OR "genomics testing" OR "genetic sequencing" OR "genetics sequencing" OR "genome sequencing" OR "genomic sequencing" OR "genomics sequencing" OR "exome sequencing" OR "exomic sequencing" OR "sequence analysis" OR "genetic profiling" OR "genome profiling" OR "genomic profiling" OR "genomics profiling" OR "direct to consumer testing" OR "direct to consumer genetic" OR "direct to consumer genetics" OR "direct to consumer genome" OR "direct to consumer genomic" OR "direct to consumer genomics" OR "personal genetic" OR "personal genetics" OR "personal genome" OR "personal genomic" OR "personal genomics" OR "individual genetic" OR "individual genetics" OR "individual genome" OR "individual genomic" OR "individual genomics" OR "genetic susceptibility testing" OR "genomic risk profiling" OR "genome based testing") *AND* TITLE: (((personal OR patient OR individual OR participant OR consumer ) AND ( utility OR meaning OR value OR benefit )))
Refined by:LANGUAGES: ( ENGLISH )

***Embase***

('genetic screening'/exp/mj OR 'genetic testing':ti OR 'genetics testing':ti OR 'genome testing':ti OR 'genomic testing':ti OR 'genomics testing':ti OR 'genetic sequencing':ti OR 'genetics sequencing':ti OR 'genome sequencing':ti OR 'genomic sequencing':ti OR 'genomics sequencing':ti OR 'exome sequencing':ti OR 'exomic sequencing':ti OR 'sequence analysis'/exp/mj OR 'sequence analysis':ti OR 'genetic profiling':ti OR 'genome profiling':ti OR 'genomic profiling':ti OR 'genomics profiling':ti OR 'direct to consumer testing':ti OR 'direct to consumer genetic':ti OR 'direct to consumer genetics':ti OR 'direct to consumer genome':ti OR 'direct to consumer genomic':ti OR 'direct to consumer genomics':ti OR 'personal genetic':ti OR 'personal genetics':ti OR 'personal genome':ti OR 'personal genomic':ti OR 'personal genomics':ti OR 'individual genetic':ti OR 'individual genetics':ti OR 'individual genome':ti OR 'individual genomic':ti OR 'individual genomics':ti OR 'genetic susceptibility testing':ti OR 'genomic risk profiling':ti OR 'genome based testing':ti) AND ('personal utility':ab,ti OR 'patient utility':ab,ti OR 'individual utility':ab,ti OR 'participant utility':ab,ti OR 'consumer utility':ab,ti OR 'public utility':ab,ti OR 'health utility':ab,ti OR 'health-related utility':ab,ti OR 'perceived utility':t,ab OR 'patient-oriented utility':ti,ab OR 'personal meaning':ab,ti OR 'patient meaning':ab,ti OR 'individual meaning':ab,ti OR 'participant meaning':ab,ti OR 'consumer meaning':ab,ti OR 'public meaning':ab,ti OR 'clinical meaning':ab,ti OR 'medical meaning':ab,ti OR 'personal value':ab,ti OR 'patient value':ab,ti OR 'individual value':ab,ti OR 'participant value':ab,ti OR 'consumer value':ab,ti OR 'public value':ab,ti OR 'psychological outcome':ab,ti OR 'psychological impact':ab,ti OR 'psychological outcomes':ab,ti OR 'psychological impacts':ab,ti OR 'personal benefit':ab,ti OR 'personal benefits':ab,ti OR 'personal harm':ab,ti OR 'personal harms':ab,ti OR 'personal outcome':ab,ti OR 'personal outcomes':ab,ti OR 'non-clinical benefit':ab,ti OR 'non-clinical benefits':ab,ti OR 'nonclinical benefit':ab,ti OR 'nonclinical benefits':ab,ti OR 'non-medical benefit':ab,ti OR 'non-medical benefits':ab,ti OR 'nonmedical benefit':ab,ti OR 'nonmedical benefits':ab,ti) AND ([english]/lim AND ([embase]/lim OR [embase classic]/lim)))

(('genetic screening'/exp/mj OR 'genetic testing':ti OR 'genetics testing':ti OR 'genome testing':ti OR 'genomic testing':ti OR 'genomics testing':ti OR 'genetic sequencing':ti OR 'genetics sequencing':ti OR 'genome sequencing':ti OR 'genomic sequencing':ti OR 'genomics sequencing':ti OR 'exome sequencing':ti OR 'exomic sequencing':ti OR 'sequence analysis'/exp/mj OR 'sequence analysis':ti OR 'genetic profiling':ti OR 'genome profiling':ti OR 'genomic profiling':ti OR 'genomics profiling':ti OR 'direct to consumer testing':ti OR 'direct to consumer genetic':ti OR 'direct to consumer genetics':ti OR 'direct to consumer genome':ti OR 'direct to consumer genomic':ti OR 'direct to consumer genomics':ti OR 'personal genetic':ti OR 'personal genetics':ti OR 'personal genome':ti OR 'personal genomic':ti OR 'personal genomics':ti OR 'individual genetic':ti OR 'individual genetics':ti OR 'individual genome':ti OR 'individual genomic':ti OR 'individual genomics':ti OR 'genetic susceptibility testing':ti OR 'genomic risk profiling':ti OR 'genome based testing':ti) AND (personal:ti OR patient:ti OR individual:ti OR participant:ti OR consumer:ti AND (utility:ti OR meaning:ti OR value:ti OR benefit:ti) AND ([english]/lim AND ([embase]/lim OR [embase classic]/lim)))

### Part B: Eligibility Criteria

Additional clarifying points regarding the eligibility criteria include:

1. Papers including various participant types (e.g., healthcare providers and patients) were included, but only data on participants classified as patients, family members of patients, or the general public were included in analysis.
2. As definitions of utility vary widely,[4] we did not limit by discipline but did require use of the term "utility." Papers broadly examining patient experiences of genetic testing were excluded;
3. Eligible studies could examine any clinical genetic or genomic test including pharmacogenomic, single gene, prenatal, exome or genome sequencing, among others. Studies of genetic testing for non-health purposes (e.g., ancestry) were not included.

### Part C: Study Screening

Our updated search identified 747 studies across the four databases. After removing 316 duplicates, 431 studies were screened for eligibility based on title and abstract content. This process identified 351 studies that were clearly ineligible. The remaining 74 studies underwent full text review by two independent reviewers, and any disagreements were reviewed and resolved by the entire study team after detailed review. Twenty-seven studies were excluded for lack of empirical data (15 studies), not directly addressing the utility (10 studies), including participants other than patients, families, or the public (1 study), or as duplicitous (1 study).

We then combined these 47 newly-identified studies with the 31 studies included in Kohler et al.'s review. We initially intended to include studies conducted in any location. However, inconsistencies in the reporting of race, ethnicity, income and (to a lesser extent) education were such that we could not be confident synthesizing the data.

We therefore excluded studies conducted in other countries after data extraction. See **Figure 1** for details.

### Part D: Data Extraction and Analysis

When extracting and synthesizing data on gender across studies, if only one category was reported (e.g., 75% women), the remaining participants were assumed to be men or women. Non-binary, transgender or queer gender identities were categorized separately when provided., Table A below provides details of the terms consolidated under each analytic category.

Table A: Gender

| | |
|---|---|
| Women | • Male<br>• Man<br>• Boy<br>• If only number of percent female was reported, the remainder were assumed male. |
| Women | • Female<br>• Woman<br>• Girl<br>• If only number of percent male was reported, the remainder were assumed female. |
| Other Gender Categories | • Nonbinary<br>• Transgender Male<br>• Transgender Female<br>• Other sex or gender |

When extracting and synthesizing data on race and ethnicity across studies, data on both race and ethnicity were recorded separately in order to incorporate all data. However, as 47 of the 54 studies included reported race and ethnicity together, our analytic categories utilize this approach in reported across all studies. When extracting data on Table B below provides details of the terms consolidated under each analytic category.

Table B: Race and Ethnicity

| | |
|---|---|
| White | • White<br>• Caucasian<br>• White/Non-Hispanic<br>• Non-Latino White<br>• White and not Hispanic or Latino<br>• Western European<br>• Other Mixed Caucasian<br>• Southern European<br>• Northern European |

| | |
|---|---|
| Black | • Black<br>• African American<br>• African American and not Hispanic or Latino<br>• Non-Hispanic Black<br>• Sub-Saharan African |
| Asian American or Pacific Islander | • Asian<br>• Asian and not Hispanic or Latino<br>• South Asian<br>• East Asian<br>• Asian American<br>• Native Hawaiian<br>• Pacific Islander<br>• Asian/Pacific Islander |
| Hispanic/Latino | • Hispanic<br>• Latinx<br>• White/Hispanic<br>• Latino<br>• Hispanic White<br>• Hispanic or Latino |
| Native American or Indigenous | • American Indian<br>• Native American<br>• Alaskan Native |
| Multiracial | • Mixed ethnicity<br>• Mixed<br>• Multiethnic/multiracial<br>• Hispanic Black<br>• Hispanic Asian |
| Other | • Other race or ethnicity<br>• Non-Hispanic or Latino, Other Race<br>• West Indian<br>• East Indian<br>• Caribbean/South American<br>• Ashkenazi Jewish |

Table C below provides details of the terms consolidated under each analytic category.

Table C: Education

| | |
|---|---|
| High School or Less | • Some primary or elementary school, or completion<br>• Some high school or secondary school, or high/secondary school degree, or GED |
| Some College | • Some community college or community college degree<br>• Vocational training |

| | |
|---|---|
| | • Two-year college or associate's degree<br>• Some technical school or technical school degree<br>• Some college, some university |
| Unspecified Less than College Degree | • Less than a college degree<br>• Greater than high school degree |
| Unspecified College Degree or Higher | • Bachelor's degree or higher<br>• College, advanced or professional degree |
| Bachelor's Degree | • College or university degree<br>• Bachelor's degree |
| Graduate Degree | • Master's degree<br>• PhD, MD, other advance coursework or postgraduate degree |

Finally, income data were typically provided as categorical data, though the income ranges used varied widely across studies. Income data were therefore consolidated into binary categories of above or below the U.S. median income of approximately $67,000. For participants in categories that crossed the median (e.g., $40,000 to $80,000), participants in these categories were distributed proportional to the population distribution within each particular household income range in the United States.[1]
