## Supplementary Appendix 4 for "Who defines the “personal utility” of genetic and genomic testing? A systematic review"

| | All Reported n(%) | Quantitative n(%) | Qualitative n(%) |
|---|---|---|---|
| **Gender** | | | |
| Women | 7,823(61.5%) | 6,573(60.2%) | 1,229(69.7%) |
| Men | 4,898(38.5%) | 4,341(39.8%) | 535(30.3%) |
| Nonbinary | 3(0.0%) | 3(0.0%) | 0(0.0%) |
| Total reported | 12,724 (100%) | 10,917 (100%) | 1,764 (100%) |
| **Race & Ethnicity**** | | | |
| White | 9,083(85.2%) | 7,950(87.7%) | 1,109(71.0%) |
| Black | 387(3.6%) | 274(3.0%) | 98(6.3%) |
| Hispanic/LatinX | 607(5.7%) | 473(5.2%) | 133(8.5%) |
| AAPI | 421(3.9%) | 382(4.2%) | 39(2.5%) |
| Native/Indigenous | 111(1.0%) | 111(1.2%) | 0(0.0%) |
| Multiracial | 27(0.3%) | 18(0.2%) | 9(0.6%) |
| Other | 412(3.9%) | 236(2.6%) | 176(11.3%) |
| Total reported | 10,663(100%) | 9,061(100%) | 1,562(100%) |
| **Education** | | | |
| Highschool or less | 899(8.2%) | 820(8.4%) | 79(6.7%) |
| Unspecified no college degree | 1,214(11.0%) | 1,062(10.9%) | 139(11.8%) |
| Some college | 1,056(9.6%) | 919(9.4%) | 137(11.6%) |
| Bachelor's degree | 893(8.1%) | 784(8.0%) | 109(9.2%) |
| Unspecified college degree | 5,497(50.0%) | 4,894(50.1%) | 576(48.8%) |
| Graduate degree | 1,440(13.1%) | 1,299(13.3%) | 141(11.9%) |
| Total reported | 10,999 (100%) | 9,778 (100%) | 1,181 (100%) |
| **Income** | | | |
| Below Median | 3,026(34.2%) | 2,764(35.0%) | 256(28.2%) |
| Above Median | 5,831(65.8%) | 5,144(65.0%) | 653(71.8%) |
| Total reported | 8,857 (100%) | 7,908 (100%) | 909 (100%) |

**Participants could select more than one category; numbers in each category exceed count for total reported
