## Supplementary Appendix 2 for "Who defines the “personal utility” of genetic and genomic testing? A systematic review"

| UPI | DOI | Title |
|---|---|---|
| 1 | 10.1002/hec.170 | Willingness-to-pay for predictive tests with no immediate treatment |
| 3 | 10.1016/j.socsci | What's at stake? Genetic information from the perspective of people |
| 5 | 10.1007/s10897- | Stakeholders' opinions on the implementation of pediatric whole exome |
| 6 | 10.1097/000020 | Reasons for seeking genetic susceptibility testing among first-degree |
| 7 | 10.1038/gim.201 | Public preferences regarding the return of individual genetic research |
| 9 | 10.1038/gim.201 | Preferences for results delivery from exome sequencing/genome |
| 10 | 10.1089/gtmb.2 | Personal factors associated with reported benefits of Huntington disease |
| 11 | 10.1089/gtmb.2 | Perception, experience, and response to genetic discrimination in |
| 13 | 10.1186/1472- | Patient and provider attitudes toward genomic testing for prostate |
| 14 | 10.1038/gim.201 | Participant use and communication of findings from exome sequencing: a |
| 15 | 10.1007/s10803- | Parents' perceptions of the usefulness of chromosomal microarray |
| 17 | 10.1002/ajmg.a. | Parental narratives on genetic testing for children with hearing loss: a |
| 18 | 10.1038/ejhg.20 | Motivators for participation in a whole-genome sequencing study: |
| 19 | 10.1038/ejhg.20 | Intentions to receive individual results from whole-genome sequencing |
| 20 | 10.1111/cge.121 | 'Information is information': a public perspective on incidental findings in |
| 21 | 10.1016/j.pec.20 | Genetic susceptibility testing for Alzheimer disease: motivation to obtain |
| 24 | 10.1111/j.1369- | BRCA mutation-negative women from hereditary breast and ovarian |
| 26 | 10.1002/hast.52 | Balancing Benefits and Risks of Immortal Data: Participants' Views of |
| 27 | 10.1002/ajmg.a. | Attitudes of non-African American focus group participants toward return |
| 28 | 10.1002/ajmg.a. | Attitudes of African Americans toward return of results from exome and |
| 29 | 10.1159/000364 | Anticipated motivation for genetic testing among smokers, nonsmokers, |
| 30 | 10.1007/s10897- | A multi-case report of the pathways to and through genetic testing and |
| 31 | 10.1097/GIM.0b | Why is genetic screening for autosomal dominant |
| 32 | 10.1038/s41436- | Parents of Newborns in the NICU Enrolled in Genome |
| 33 | https://doi.org/1 | A Prospective Study of Parental Perceptions |
| 35 | 10.1002/jgc4.11 | The personal utility of cfDNA screening: Pregnant patients' |
| 36 | 62619 | Perceived utility and disutility of genomic sequencing for |
| 37 | 10.1002/jgc4.13 | Comprehension and personal value of negative non-diagnostic |
| 38 | 10.1007/s12687- | Great expectations: patient perspectives and anticipated utility |
| 41 | 10.1111/cge.129 | Defining personal utility in genomics: A Delphi study |
| 43 | 10.2217/pme.15 | Patients' perceived utility of |
| 44 | https://doi.org/1 | Parental Perspectives on |
| 45 | 10.1001/jamaps | Identification of Neuropsychiatric Copy Number Variants |
| 48 | https://doi.org/1 | Perceived Benefits, Risks, and |
| 49 | 10.1007/s10897- | "Is It Worth Knowing?" Focus Group Participants' Perceived |
| 50 | https://doi.org/1 | Perceived Utility of Genomic Sequencing: Qualitative Analysis |
| 53 | https://doi.org/1 | Population Genomic Screening for Genetic Etiologies of |
| 55 | 10.1186/s12884- | The influence of experiential knowledge |
| 56 | 10.1002/jgc4.10 | Much Ado about Nothing: a Qualitative Study of the Experiences |
| 58 | 10.1038/s41436- | Parents' perceptions of personal utility of exome sequencing results |
| 60 | 10.1200/PO.17.0 | Parental Perspectives on Whole Exome Sequencing in Pediatric Cancer: A |
| 61 | 10.1177/000992 | Parental Perception and Participation in Genetic Testing Among Children |
| 62 | 017-0090-y | act of Panel Gene Testing for Hereditary Breast and Ovarian Cancer on Pati |
| 64 | 7.223 | atisfaction with, and perceived utility of whole-genome sequencing: finding |
| 67 | 018-0344-6 | ers' views on the value of outcomes from clinical genetic and genomic inte |

| | | |
|---|---|---|
| 68 | 019-0619-9 | cing in healthy adults: design, participant characteristics, and early outcom |
| 70 | 60 | vith sporadic and familial amyotrophic lateral sclerosis found value in gene |
| 71 | 020.0275 | comes and Experiences with Population Genetic Testing Offered Through a |
| 72 | 018-0161-z | massively parallel sequencing genetic testing of colorectal cancer risk: a dis |
| 75 | 006 | umer Genetic Testing: User Motivations, Decision Making, and Perceived U |
| 76 | 468 | er genetic testing for factor V Leiden and prothrombin 20210G>A: the cons |
| 77 | 018-0321-0 | l utility of expanded carrier screening: results-guided actionability and out |
| 79 | 6.194 | d screening for fetal aneuploidy by primary obstetrical care providers in the |

| Journal | Year | N_Study | N_Patient | N_Family | N_Public | N_IneligIble |
|---|---|---|---|---|---|---|
| Health Econ. | 2012 | 1463 | 0 | 0 | 1463 | 0 |
| Soc Sci Med. | 2011 | 40 | 22 | 18 | 0 | 0 |
| Journal of Genetic Counseling | 2014 | 49 | 7 | 20 | 0 | 22 |
| Alzheimer Disease and Associated | 2003 | 206 | 0 | 206 | 0 | 0 |
| Genetics in Medicine: Official | 2012 | 89 | 0 | 0 | 89 | 0 |
| Genetics in Medicine: Official | 2014 | 39 | 39 | 0 | 0 | 0 |
| Genetic Testing and Molecular | 2010 | 74 | 74 | 0 | 0 | 0 |
| Genetic Testing and Molecular | 2013 | 60 | 60 | 0 | 0 | 0 |
| BMC health services research | 2013 | 47 | 0 | 23 | 0 | 24 |
| Genetics in Medicine: Official | 2016 | 29 | 29 | 0 | 0 | 0 |
| Journal of Autism and | 2015 | 57 | 0 | 57 | 0 | 0 |
| American Journal of Medical | 2007 | 18 | 0 | 18 | 0 | 0 |
| European journal of human | 2011 | 322 | 322 | 0 | 0 | 0 |
| European journal of human | 2013 | 311 | 311 | 0 | 0 | 0 |
| Clinical Genetics | 2013 | 63 | 0 | 0 | 63 | 0 |
| Patient Education and Counseling | 2006 | 60 | 0 | 60 | 0 | 0 |
| Health Expectations: An | 2008 | 13 | 13 | 0 | 0 | 0 |
| The Hastings Center Report | 2016 | 34 | 0 | 0 | 34 | 0 |
| American Journal of Medical | 2014 | 35 | 0 | 35 | 0 | 0 |
| American Journal of Medical | 2013 | 41 | 0 | 41 | 0 | 0 |
| Public Health Genomics | 2014 | 755 | 0 | 0 | 755 | 0 |
| Journal of Genetic Counseling | 2013 | 3 | 3 | 0 | 0 | 0 |
| Genetics IN Medicine | 2011 | 119 | 109 | 10 | 0 | 0 |
| Genet Med | 2020 | 23 | 0 | 23 | 0 | 0 |
| e American Journal of Human Genet | 2020 | 312 | 0 | 312 | 0 | 0 |
| Journal of Genetic Counseling | 2019 | 27 | 0 | 27 | 0 | 0 |
| merican Journal of Medical Genetic | 2021 | 30 | 0 | 30 | 0 | 0 |
| Journal of Genetic Counseling | 2021 | 17 | 17 | 0 | 0 | 0 |
| J Community Genet | 2018 | 14 | 14 | 0 | 0 | 0 |
| Clin Genet | 2017 | 40 | 40 | 0 | 0 | 0 |
| Personalized Medicine | 2016 | 202 | 202 | 0 | 0 | 0 |
| Precision Oncology | 2017 | 64 | 0 | 64 | 0 | 0 |
| JAMA Psychiatry | 2020 | 141 | 141 | 0 | 0 | 0 |
| Pediatrics | 2019 | 493 | 0 | 493 | 0 | 0 |
| Journal of Genetic Counseling | 2016 | 16 | 16 | 0 | 0 | 0 |
| ient - Patient-Centered Outcomes R | 2021 | 60 | 20 | 40 | 0 | 0 |
| Journal of Personalized Medicine | 2021 | 27 | 27 | 0 | 0 | 0 |
| BMC Pregnancy and Childbirth | 2020 | 25 | 25 | 0 | 0 | 0 |
| Journal of Genetic Counseling | 2019 | 12 | 12 | 0 | 0 | 0 |
| Genetics in Medicine: Official | 2020 | 31 | 0 | 31 | 0 | 0 |
| JCO precision oncology | 2017 | 64 | 0 | 64 | 0 | 0 |
| Clinical Pediatrics | 2018 | 143 | 0 | 143 | 0 | 0 |
| J Genet Couns | 2017 | 232 | 232 | 0 | 0 | 0 |
| Genet Med | 2018 | 202 | 202 | 0 | 0 | 0 |
| Genet Med | 2019 | 126 | 5 | 1 | 0 | 120 |

| | | | | | | |
|---|---|---|---|---|---|---|
| Genome Med | 2019 | 543 | 0 | 0 | 543 | 0 |
| Mol. Genetics Genom. Med. | 2018 | 449 | 449 | 0 | 0 | 0 |
| Genet Test Mol Biomarkers | 2021 | 1646 | 1646 | 0 | 0 | 0 |
| Eur J Hum Genet | 2018 | 122 | 122 | 0 | 0 | 0 |
| Public Health Genomics | 2017 | 1648 | 0 | 0 | 1648 | 0 |
| Mol Genet Genomic Med | 2020 | 2354 | 1244 | 0 | 1110 | 0 |
| Genet Med | 2019 | 391 | 391 | 0 | 0 | 0 |
| Genet Med | 2017 | 100 | 100 | 0 | 0 | 0 |

| N_Eligible | Method_Type | Edu_Rep | N_HS_OrLess | N_Some_Colleg | N_LessThan_Col |
|---|---|---|---|---|---|
| 1463 | Qual | Yes | 585 | 424 | 0 |
| 40 | Quant | Yes | 3 | 10 | 0 |
| 27 | Quant | Yes | 2 | 4 | 0 |
| 206 | Qual | Yes | 0 | 0 | 0 |
| 89 | Quant | No | 0 | 0 | 0 |
| 39 | Quant | No | 0 | 0 | 0 |
| 74 | Qual | No | 0 | 0 | 0 |
| 60 | Qual | Yes | 21 | 19 | 0 |
| 23 | Quant | Yes | 2 | 9 | 0 |
| 29 | Quant | Yes | 1 | 5 | 0 |
| 57 | Quant | Yes | 12 | 14 | 0 |
| 18 | Quant | Yes | 6 | 2 | 0 |
| 322 | Qual | Yes | 8 | 42 | 0 |
| 311 | Qual | Yes | 0 | 0 | 49 |
| 63 | Quant | Yes | 24 | 13 | 0 |
| 60 | Quant | Yes | 0 | 0 | 0 |
| 13 | Quant | Yes | 0 | 0 | 0 |
| 34 | Quant | No | 0 | 0 | 0 |
| 35 | Quant | Yes | 0 | 0 | 14 |
| 41 | Quant | Yes | 0 | 0 | 31 |
| 755 | Qual | Yes | 0 | 0 | 183 |
| 3 | Quant | No | 0 | 0 | 0 |
| 119 | Quant | Yes | 9 | 37 | 0 |
| 23 | Quant | Yes | 0 | 0 | 9 |
| 312 | Qual | Yes | 142 | 0 | 0 |
| 27 | Quant | Yes | 0 | 5 | 0 |
| 30 | Quant | Yes | 11 | 3 | 0 |
| 17 | Quant | Yes | 0 | 3 | 0 |
| 14 | Quant | Yes | 0 | 2 | 0 |
| 40 | Other | Yes | 0 | 0 | 13 |
| 202 | Qual | Yes | 0 | 0 | 38 |
| 64 | Quant | Yes | 0 | 0 | 13 |
| 141 | Quant | No | 0 | 0 | 0 |
| 493 | Quant | Yes | 0 | 0 | 59 |
| 16 | Quant | No | 0 | 0 | 0 |
| 60 | Quant | Yes | 8 | 12 | 0 |
| 27 | Quant | No | 0 | 0 | 0 |
| 25 | Quant | No | 0 | 0 | 0 |
| 12 | Quant | Yes | 0 | 2 | 0 |
| 31 | Quant | Yes | 1 | 16 | 0 |
| 64 | Quant | Yes | 0 | 0 | 13 |
| 143 | Quant | No | 0 | 0 | 0 |
| 232 | Qual | Yes | 0 | 0 | 63 |
| 202 | Qual | Yes | 0 | 0 | 38 |
| 6 | Other | No | 0 | 0 | 0 |

| | | | | | |
|---|---|---|---|---|---|
| 543 | Qual | Yes | 0 | 0 | 24 |
| 449 | Qual | No | 0 | 0 | 0 |
| 1646 | Qual | Yes | 0 | 0 | 304 |
| 122 | Qual | Yes | 13 | 80 | 0 |
| 1648 | Qual | Yes | 0 | 0 | 363 |
| 2354 | Qual | Yes | 51 | 354 | 0 |
| 391 | Qual | No | 0 | 0 | 0 |
| 100 | Qual | No | 0 | 0 | 0 |

| N_CollegeDeg_O | N_Bachelor_Deg | N_Graduate_De | N_Edu_Miss | N_Edu_Rep_Tot | N_ Inc_Rep |
|---|---|---|---|---|---|
| 454 | 0 | 0 | 0 | 1463 | Yes |
| 0 | 14 | 11 | 2 | 38 | Yes |
| 0 | 0 | 14 | 7 | 20 | No |
| 0 | 0 | 0 | 206 | 0 | Yes |
| 0 | 0 | 0 | 89 | 0 | No |
| 0 | 0 | 0 | 39 | 0 | Yes |
| 0 | 0 | 0 | 74 | 0 | No |
| 0 | 3 | 9 | 8 | 52 | No |
| 0 | 7 | 5 | 0 | 23 | Yes |
| 0 | 6 | 16 | 1 | 28 | No |
| 0 | 16 | 15 | 0 | 57 | No |
| 0 | 5 | 5 | 0 | 18 | Yes |
| 0 | 98 | 168 | 6 | 316 | Yes |
| 253 | 0 | 0 | 9 | 302 | Yes |
| 0 | 11 | 15 | 0 | 63 | No |
| 0 | 0 | 0 | 60 | 0 | Yes |
| 0 | 0 | 0 | 13 | 0 | No |
| 0 | 0 | 0 | 34 | 0 | No |
| 21 | 0 | 0 | 0 | 35 | Yes |
| 10 | 0 | 0 | 0 | 41 | Yes |
| 572 | 0 | 0 | 0 | 755 | No |
| 0 | 0 | 0 | 3 | 0 | No |
| 73 | 0 | 0 | 0 | 119 | No |
| 14 | 0 | 0 | 0 | 23 | No |
| 0 | 88 | 82 | 0 | 312 | No |
| 0 | 10 | 12 | 0 | 27 | No |
| 0 | 7 | 9 | 0 | 30 | Yes |
| 0 | 7 | 7 | 0 | 17 | Yes |
| 0 | 7 | 5 | 0 | 14 | Yes |
| 27 | 0 | 0 | 0 | 40 | Yes |
| 164 | 0 | 0 | 0 | 202 | Yes |
| 12 | 0 | 0 | 39 | 25 | Yes |
| 0 | 0 | 0 | 141 | 0 | No |
| 434 | 0 | 0 | 0 | 493 | Yes |
| 0 | 0 | 0 | 16 | 0 | No |
| 0 | 10 | 12 | 18 | 42 | No |
| 0 | 0 | 0 | 27 | 0 | No |
| 0 | 0 | 0 | 25 | 0 | Yes |
| 0 | 4 | 6 | 0 | 12 | No |
| 0 | 5 | 9 | 0 | 31 | No |
| 12 | 0 | 0 | 39 | 25 | Yes |
| 0 | 0 | 0 | 143 | 0 | Yes |
| 169 | 0 | 0 | 0 | 232 | No |
| 164 | 0 | 0 | 0 | 202 | Yes |
| 0 | 0 | 0 | 6 | 0 | No |

| | | | | | |
|---|---|---|---|---|---|
| 505 | 0 | 0 | 14 | 529 | Yes |
| 0 | 0 | 0 | 449 | 0 | No |
| 1329 | 0 | 0 | 13 | 1633 | Yes |
| 0 | 0 | 29 | 0 | 122 | Yes |
| 1284 | 0 | 0 | 1 | 1647 | Yes |
| 0 | 595 | 1011 | 343 | 2011 | Yes |
| 0 | 0 | 0 | 391 | 0 | No |
| 0 | 0 | 0 | 100 | 0 | No |

| N_Inc_Miss | N_Inc_BelowMe | N_Inc_AboveMe | N_Inc_Rep | Gen_Rep | Gen_Women_O |
|---|---|---|---|---|---|
| 0 | 891 | 572 | 1463 | Yes | 0 |
| 4 | 16 | 20 | 36 | Yes | 0 |
| 27 | 0 | 0 | 0 | Yes | 0 |
| 206 | 0 | 0 | 0 | Yes | 0 |
| 89 | 0 | 0 | 0 | No | 0 |
| 0 | 5 | 34 | 39 | Yes | 0 |
| 74 | 0 | 0 | 0 | Yes | 0 |
| 60 | 0 | 0 | 0 | Yes | 0 |
| 3 | 7 | 13 | 20 | No | 0 |
| 29 | 0 | 0 | 0 | Yes | 0 |
| 57 | 0 | 0 | 0 | Yes | 0 |
| 0 | 10 | 8 | 18 | Yes | 0 |
| 18 | 35 | 269 | 304 | Yes | 0 |
| 16 | 52 | 243 | 295 | Yes | 0 |
| 63 | 0 | 0 | 0 | Yes | 0 |
| 60 | 0 | 0 | 0 | Yes | 0 |
| 13 | 0 | 0 | 0 | Yes | 1 |
| 34 | 0 | 0 | 0 | Yes | 0 |
| 0 | 17 | 18 | 35 | Yes | 0 |
| 0 | 31 | 10 | 41 | Yes | 0 |
| 755 | 0 | 0 | 0 | Yes | 0 |
| 3 | 0 | 0 | 0 | Yes | 1 |
| 119 | 0 | 0 | 0 | Yes | 0 |
| 23 | 0 | 0 | 0 | Yes | 0 |
| 312 | 0 | 0 | 0 | Yes | 0 |
| 27 | 0 | 0 | 0 | Yes | 1 |
| 0 | 9 | 21 | 30 | Yes | 0 |
| 0 | 8 | 9 | 17 | Yes | 0 |
| 1 | 1 | 12 | 13 | Yes | 0 |
| 0 | 6 | 34 | 40 | Yes | 0 |
| 7 | 55 | 140 | 195 | Yes | 0 |
| 64 | 0 | 0 | 0 | Yes | 0 |
| 141 | 0 | 0 | 0 | Yes | 0 |
| 10 | 75 | 408 | 483 | Yes | 0 |
| 16 | 0 | 0 | 0 | Yes | 1 |
| 60 | 0 | 0 | 0 | Yes | 0 |
| 27 | 0 | 0 | 0 | Yes | 0 |
| 0 | 2 | 23 | 25 | Yes | 1 |
| 12 | 0 | 0 | 0 | Yes | 0 |
| 31 | 0 | 0 | 0 | Yes | 0 |
| 43 | 13 | 8 | 21 | Yes | 0 |
| 12 | 62 | 69 | 131 | Yes | 0 |
| 232 | 0 | 0 | 0 | Yes | 0 |
| 28 | 34 | 140 | 174 | Yes | 0 |
| 6 | 0 | 0 | 0 | No | 0 |

| | | | | | |
|---|---|---|---|---|---|
| 28 | 91 | 424 | 515 | Yes | 0 |
| 449 | 0 | 0 | 0 | No | 0 |
| 109 | 420 | 1117 | 1537 | Yes | 0 |
| 32 | 51 | 39 | 90 | Yes | 0 |
| 25 | 695 | 928 | 1623 | Yes | 0 |
| 642 | 440 | 1272 | 1712 | Yes | 0 |
| 391 | 0 | 0 | 0 | Yes | 1 |
| 100 | 0 | 0 | 0 | Yes | 1 |

| N_Women | N_Men | N_Gen_Other | N_Gen_Miss | N_Gen_Rep_Tot | Race_Rep |
|---|---|---|---|---|---|
| 717 | 746 | 0 | 0 | 1463 | No |
| 28 | 12 | 0 | 0 | 40 | No |
| 22 | 5 | 0 | 0 | 27 | Yes |
| 149 | 57 | 0 | 0 | 206 | Yes |
| 0 | 0 | 0 | 89 | 0 | Yes |
| 16 | 23 | 0 | 0 | 39 | Yes |
| 54 | 20 | 0 | 0 | 74 | No |
| 39 | 21 | 0 | 0 | 60 | No |
| 0 | 0 | 0 | 23 | 0 | Yes |
| 14 | 15 | 0 | 0 | 29 | Yes |
| 48 | 9 | 0 | 0 | 57 | Yes |
| 14 | 4 | 0 | 0 | 18 | Yes |
| 182 | 140 | 0 | 0 | 322 | Yes |
| 128 | 183 | 0 | 0 | 311 | Yes |
| 45 | 18 | 0 | 0 | 63 | Yes |
| 52 | 8 | 0 | 0 | 60 | Yes |
| 13 | 0 | 0 | 0 | 13 | Yes |
| 14 | 20 | 0 | 0 | 34 | Yes |
| 29 | 6 | 0 | 0 | 35 | Yes |
| 29 | 12 | 0 | 0 | 41 | Yes |
| 542 | 213 | 0 | 0 | 755 | Yes |
| 3 | 0 | 0 | 0 | 3 | No |
| 87 | 32 | 0 | 0 | 119 | Yes |
| 17 | 6 | 0 | 0 | 23 | Yes |
| 170 | 142 | 0 | 0 | 312 | Yes |
| 27 | 0 | 0 | 0 | 27 | Yes |
| 27 | 3 | 0 | 0 | 30 | Yes |
| 14 | 3 | 0 | 0 | 17 | Yes |
| 5 | 9 | 0 | 0 | 14 | Yes |
| 18 | 22 | 0 | 0 | 40 | Yes |
| 101 | 101 | 0 | 0 | 202 | Yes |
| 56 | 8 | 0 | 0 | 64 | Yes |
| 95 | 46 | 0 | 0 | 141 | No |
| 246 | 247 | 0 | 0 | 493 | Yes |
| 15 | 1 | 0 | 0 | 16 | Yes |
| 49 | 11 | 0 | 0 | 60 | Yes |
| 17 | 10 | 0 | 0 | 27 | No |
| 25 | 0 | 0 | 0 | 25 | Yes |
| 4 | 8 | 0 | 0 | 12 | Yes |
| 27 | 4 | 0 | 0 | 31 | Yes |
| 56 | 8 | 0 | 0 | 64 | Yes |
| 135 | 7 | 0 | 1 | 142 | Yes |
| 224 | 8 | 0 | 0 | 232 | Yes |
| 101 | 101 | 0 | 0 | 202 | Yes |
| 3 | 0 | 0 | 3 | 3 | No |

| | | | | | |
|---|---|---|---|---|---|
| 202 | 326 | 2 | 13 | 530 | Yes |
| 0 | 0 | 0 | 449 | 0 | No |
| 1152 | 480 | 1 | 13 | 1633 | Yes |
| 65 | 57 | 0 | 0 | 122 | No |
| 995 | 653 | 0 | 0 | 1648 | Yes |
| 1261 | 1093 | 0 | 0 | 2354 | Yes |
| 391 | 0 | 0 | 0 | 391 | Yes |
| 100 | 0 | 0 | 0 | 100 | No |

| Race_Rep_Notes | Race_Focus | N_NHWhite | N_Black |
|---|---|---|---|
| | No | 0 | 0 |
| | No | 0 | 0 |
| lispanic White and Hisp | No | 25 | 0 |
| Only White | No | 195 | 0 |
| | No | 0 | 0 |
| Only White and Asian | No | 34 | 0 |
| | No | 0 | 0 |
| | No | 0 | 0 |
| | No | 17 | 3 |
| | No | 25 | 1 |
| | No | 47 | 6 |
| | No | 12 | 1 |
| | No | 286 | 13 |
| | No | 253 | 16 |
| | No | 42 | 10 |
| Only White | No | 57 | 0 |
| Only White | No | 10 | 0 |
| | No | 32 | 0 |
| | No | 31 | 0 |
| | Yes | 0 | 41 |
| | No | 650 | 27 |
| | No | 0 | 0 |
| Only White | No | 110 | 0 |
| | No | 19 | 0 |
| | No | 129 | 7 |
| | No | 25 | 1 |
| | No | 11 | 0 |
| | No | 7 | 5 |
| | No | 8 | 1 |
| | No | 24 | 15 |
| Only White | No | 177 | 0 |
| Only White and Hispanic | No | 25 | 0 |
| | No | 0 | 0 |
| Only White and Hispanic | No | 335 | 0 |
| nority participants, all p | No | 16 | 0 |
| | No | 28 | 13 |
| | No | 0 | 0 |
| Only White | No | 24 | 0 |
| | No | 9 | 0 |
| | No | 28 | 1 |
| | No | 25 | 0 |
| | No | 107 | 15 |
| | No | 138 | 23 |
| Only White | No | 176 | 0 |
| | No | 0 | 0 |

| | | | |
|---|---|---|---|
| | No | 485 | 3 |
| | No | 0 | 0 |
| | No | 1443 | 49 |
| | No | 0 | 0 |
| | No | 1484 | 66 |
| | No | 2158 | 56 |
| | No | 376 | 14 |
| | No | 0 | 0 |

| N_HispLatx | N_AAPI | N_Native_Indig | N_Multiracial | N_Race_Other |
|---|---|---|---|---|
| 0 | 0 | 0 | 0 | 0 |
| 0 | 0 | 0 | 0 | 0 |
| 0 | 0 | 0 | 0 | 0 |
| 0 | 0 | 0 | 0 | 0 |
| 0 | 0 | 0 | 0 | 0 |
| 0 | 5 | 0 | 0 | 0 |
| 0 | 0 | 0 | 0 | 0 |
| 0 | 0 | 0 | 0 | 0 |
| 3 | 0 | 0 | 0 | 0 |
| 0 | 3 | 0 | 0 | 0 |
| 0 | 1 | 0 | 3 | 0 |
| 4 | 1 | 0 | 0 | 0 |
| 6 | 15 | 0 | 0 | 6 |
| 0 | 19 | 0 | 0 | 0 |
| 6 | 5 | 0 | 0 | 0 |
| 0 | 0 | 0 | 0 | 0 |
| 0 | 0 | 0 | 0 | 3 |
| 0 | 1 | 0 | 1 | 0 |
| 0 | 3 | 0 | 0 | 1 |
| 0 | 0 | 0 | 0 | 0 |
| 22 | 34 | 0 | 18 | 0 |
| 0 | 0 | 0 | 0 | 0 |
| 0 | 0 | 0 | 0 | 0 |
| 2 | 0 | 0 | 0 | 4 |
| 120 | 20 | 2 | 0 | 24 |
| 1 | 0 | 0 | 0 | 0 |
| 13 | 4 | 0 | 2 | 0 |
| 5 | 0 | 0 | 0 | 0 |
| 3 | 2 | 0 | 0 | 0 |
| 1 | 0 | 0 | 0 | 0 |
| 0 | 0 | 0 | 0 | 25 |
| 27 | 0 | 0 | 0 | 12 |
| 0 | 0 | 0 | 0 | 0 |
| 32 | 0 | 0 | 0 | 126 |
| 0 | 0 | 0 | 0 | 0 |
| 7 | 9 | 0 | 3 | 0 |
| 0 | 0 | 0 | 0 | 0 |
| 0 | 0 | 0 | 0 | 0 |
| 1 | 2 | 0 | 0 | 0 |
| 0 | 2 | 0 | 0 | |
| 27 | 0 | 0 | 0 | 12 |
| 2 | 1 | 0 | 0 | 18 |
| 38 | 15 | 0 | 0 | 16 |
| 0 | 0 | 0 | 0 | 26 |
| 0 | 0 | 0 | 0 | 0 |

| | | | | |
|---|---|---|---|---|
| 16 | 15 | 0 | 0 | 26 |
| 0 | 0 | 0 | 0 | 0 |
| 85 | 146 | 0 | 0 | 0 |
| 0 | 0 | 0 | 0 | 0 |
| 88 | 0 | 0 | 0 | 10 |
| 79 | 57 | 106 | 0 | 91 |
| 19 | 61 | 3 | 0 | 12 |
| 0 | 0 | 0 | 0 | 0 |

| N_Race_Miss | N_Race_Rep_To | Race_Discrete |
|---|---|---|
| 1463 | 0 | Yes |
| 40 | 0 | Yes |
| 2 | 25 | Yes |
| 11 | 195 | Yes |
| 89 | 0 | Yes |
| 0 | 39 | Yes |
| 74 | 0 | Yes |
| 60 | 0 | Yes |
| 0 | 23 | Yes |
| 0 | 29 | Yes |
| 0 | 57 | Yes |
| 0 | 18 | Yes |
| 0 | 326 | No |
| 23 | 288 | Yes |
| 0 | 63 | Yes |
| 3 | 57 | Yes |
| 0 | 13 | Yes |
| 0 | 34 | Yes |
| 0 | 35 | Yes |
| 0 | 41 | Yes |
| 4 | 751 | Yes |
| 3 | 0 | Yes |
| 9 | 110 | Yes |
| 0 | 25 | No |
| 10 | 302 | Yes |
| 0 | 27 | Yes |
| 0 | 30 | Yes |
| 0 | 17 | Yes |
| 0 | 14 | Yes |
| 0 | 40 | Yes |
| 0 | 202 | Yes |
| 0 | 64 | Yes |
| 141 | 0 | Yes |
| 0 | 493 | Yes |
| 0 | 16 | Yes |
| 0 | 60 | Yes |
| 27 | 0 | Yes |
| 1 | 24 | Yes |
| 0 | 12 | Yes |
| 0 | 31 | Yes |
| 0 | 64 | Yes |
| 0 | 143 | Yes |
| 2 | 230 | Yes |
| 0 | 202 | Yes |
| 6 | 0 | Yes |

| | | |
|---|---|---|
| 0 | 545 | No |
| 449 | 0 | Yes |
| 0 | 1723 | No |
| 122 | 0 | Yes |
| 0 | 1648 | Yes |
| 0 | 2547 | No |
| 13 | 485 | No |
| 100 | 0 | Yes |
